# Prevalence, Demographic Patterns, and Seasonal Distribution of Malaria in District Dera Ismail Khan, Pakistan

**DOI:** 10.64898/2025.12.29.25343127

**Authors:** Faiqa Fateen, Abdus Sami, Sadaf fatimah, Hugo Douma, Momna Elham

## Abstract

Malaria remains the most significant vector-borne disease worldwide, with over 200 million cases reported annually, causing approximately 0.6 million deaths among children and pregnant women. In Pakistan, particularly in the southern regions of Khyber Pakhtunkhwa, malaria continues to pose a major public health challenge. This study aimed to determine the monthly prevalence of malaria parasites in the population of Dera Ismail Khan and to identify the patterns of parasite transmission. A retrospective analysis was conducted using laboratory-confirmed malaria case data obtained from the District Health Office (DHO) Dera Ismail Khan for the entire year of 2024. Cases were categorized by month and Plasmodium species, specifically *P. vivax, P. falciparum*, and mixed infections. Chi-square analysis was performed to assess the significance of monthly variations in species distribution. The results showed that *P. vivax* was the predominant species, with cases peaking from September to December. *P. falciparum* and mixed infections were comparatively rare. October recorded the highest number of cases, followed by September and November. Chi-square analysis confirmed a statistically significant association between month and species distribution (χ^2^ = 2314.40, df = 22, p < 0.0001), indicating a strong seasonal pattern. Malaria incidence was higher in males than females, and the majority of cases occurred among individuals aged 15–24 years. Overall, the study demonstrates that *P. vivax* is the most common malaria parasite in Dera Ismail Khan and that malaria transmission in the region exhibits a clear seasonal trend. These findings provide valuable insight for targeted malaria control and prevention strategies in endemic areas of Khyber Pakhtunkhwa.

**Author Summary:** Malaria remains a major public health problem worldwide, but not all malaria parasites behave in the same way. In this study, we focused on malaria in District Dera Ismail Khan, Pakistan, with particular attention to *Plasmodium vivax*, a species that has historically received less attention than *Plasmodium falciparum*. Using routine health records from 2024, we analyzed nearly 37,000 laboratory-confirmed malaria cases to understand which parasite species were most common and how infections varied by age, sex, and season.

We found that *Plasmodium vivax* caused more than four out of five malaria cases in the district, making it the dominant species across all age groups. Malaria was more common in males than females, and clear seasonal patterns were observed, with most cases occurring after the monsoon season between September and November. These findings are important because *P. vivax* can cause repeated illness through relapses, making malaria harder to control even when mosquito numbers are low.

By documenting local patterns of malaria transmission, our study highlights the need for targeted control strategies that address *P. vivax* specifically. This information can help public health authorities plan more effective, seasonally timed interventions and move closer to malaria elimination.

## Introduction

The two Plasmodium species associated with malaria in humans are *Plasmodium vivax* and *Plasmodium falciparum*. Until recent years, most malaria research and funding focused primarily on the prevention, treatment, and control of *P. falciparum* (1). Both parasites place approximately 2.5 billion individuals at risk. Although *P. falciparum* is less prevalent than *P. vivax* in sub-Saharan Africa, it is responsible for a higher number of deaths. In many densely populated and impoverished regions, *P. vivax* predominates, and recent evidence linking it to severe and lethal outcomes highlights the need to address this historically neglected infection. Effective malaria control and elimination require a deeper understanding of the unique epidemiological features of this parasite (2).

This review is therefore limited to such a synthesis of current knowledge. Significant differences in the biology of *P. vivax* and *P. falciparum* result in distinct epidemiological patterns. Notably, *P. vivax* can cause relapses weeks to months after an initial infection through activation of dormant liver-stage parasites called hypnozoites. This dormant liver-stage infection, known as the hypnozoite reservoir, generates new blood-stage infections and clinical attacks within affected communities, contributing to further transmission. Hypnozoite-driven infections can occur even in seasons when the local *Anopheles* mosquito is less active, effectively expanding *P. vivax*’s geographic range into temperate areas, such as the Korean Peninsula (3).

In humans, five *Plasmodium* species cause malaria: *P. vivax, P. falciparum, P. ovale, P. malariae*, and *P. knowlesi* (a zoonotic infection). Despite extensive and costly malaria control efforts over decades, malaria has re-emerged as a major public health issue in Asia. In Pakistan, 60 percent of the population resides in malaria-endemic areas. *P. vivax* accounts for 81.3 percent of cases, *P. falciparum* 14.7 percent, and mixed infections 4 percent (4).

Globally, malaria remains the most prevalent vector-borne disease, causing over 200 million clinical cases annually. Approximately 0.6 million child and maternal deaths are attributed to malaria each year. Current trends indicate that the world is not on track to meet the 2020 milestones of the World Health Organization (WHO) Global Technical Strategy (GTS) for malaria 2016– 2030, which aims to reduce malaria-related deaths and cases by 40 percent by 2020. Malaria incidence has steadily increased from 2015 to 2018, with 214 million, 217 million, and 219 million cases reported in 2015, 2016, and 2017, respectively (5).

Pakistan has a tropical to temperate climate, with arid conditions along the southern coast and altitudes ranging from sea level to nearly 9,000 meters (6). The primary *Plasmodium* species in Pakistan are *P. vivax* (causing approximately 64 percent of cases) and *P. falciparum* (causing roughly 36 percent of cases), with malaria predominantly reported in Khyber Pakhtunkhwa, Baluchistan, Sindh, and the Federally Administered Tribal Areas (7). Malaria transmission exhibits seasonal variations: *P. vivax* transmission peaks between June and September, with relapses from previous seasons occurring from April to June, whereas *P. falciparum* transmission peaks from August to December (8).

The WHO classifies Pakistan as a country with moderate malaria prevalence that has implemented substantial malaria control programs. Nevertheless, estimates indicate that about 50 deaths occur per 500,000 reported malaria cases annually (IRIN = Integrated Regional Information Networks, 2007) (9).

## Materials and Methods

### Study Area Description

District Dera Ismail Khan (D.I. Khan) is a prominent city in the Khyber Pakhtunkhwa province of Pakistan, noted for its aesthetic appeal. The district is geographically located at a latitude of 31.8224° N and a longitude of 70.8940° E. It is bordered by Dera Ghazi Khan and Muzaffargarh to the east, Bannu District to the north, the Sulaiman Range to the south, and South Waziristan to the west. According to the 2023 census reported by Google, the population of Dera Ismail Khan District stands at approximately 1,900,000.

### Study Design and Setting

This study employed a cross-sectional, retrospective design to determine the prevalence and demographic patterns of malaria in Dera Ismail Khan District, Khyber Pakhtunkhwa, Pakistan. The district is recognized as endemic for malaria, and health surveillance data are routinely collected and maintained by the Office of the District Health Officer (DHO). This setting provided a representative sample of the local population for analysis.

### Data Source and Population

Data were obtained from official DHO records, which included laboratory-confirmed malaria cases reported between January and December 2024. The dataset consisted of de-identified patient-level information, including sex, age, and *Plasmodium* species. Only confirmed infections of *Plasmodium vivax, P. falciparum*, and mixed infections were included in the analysis. Cases that were incomplete or lacked species identification were excluded.

### Statistical Analysis

Descriptive statistics were used to present the prevalence of each malaria type by age and sex. The Chi-square (χ^2^) test of independence was applied to examine associations between categorical variables, such as species versus sex or species versus age group. Expected frequencies were calculated under the assumption of no relationship between variables. Statistical significance was defined as p < 0.05 with a 95% confidence interval. Microsoft Excel was used for all statistical analyses.

### Ethical Considerations

As the study utilized secondary data from public health records, there was no direct contact with patients or any intervention. All personally identifiable information was excluded to ensure confidentiality. Since the data were anonymized and aggregated, institutional ethical clearance was not required.

## Results

### Species-wise Prevalence of Malaria

A total of 36,938 malaria cases were recorded in the study. Of these, 29,718 cases (80.46%) were caused by *Plasmodium vivax*, 7,160 cases (19.39%) by *P. falciparum*, and only 60 cases (0.16%) represented mixed infections. The results indicate that *P. vivax* was substantially more prevalent, while mixed infections were rare (Table 1) (Figure 1).

**Table 1:** Distribution of Malarial Parasites Among Infected Population.

| Species | Observed |
| --- | --- |
| *P. vivax* | 29718 |
| *P. falciparum* | 7160 |
| Mixed | 60 |
| $\chi^2 = 38,274.97$; $df = 2$; $\alpha = 5.99$ | |

**Fig. 1:**
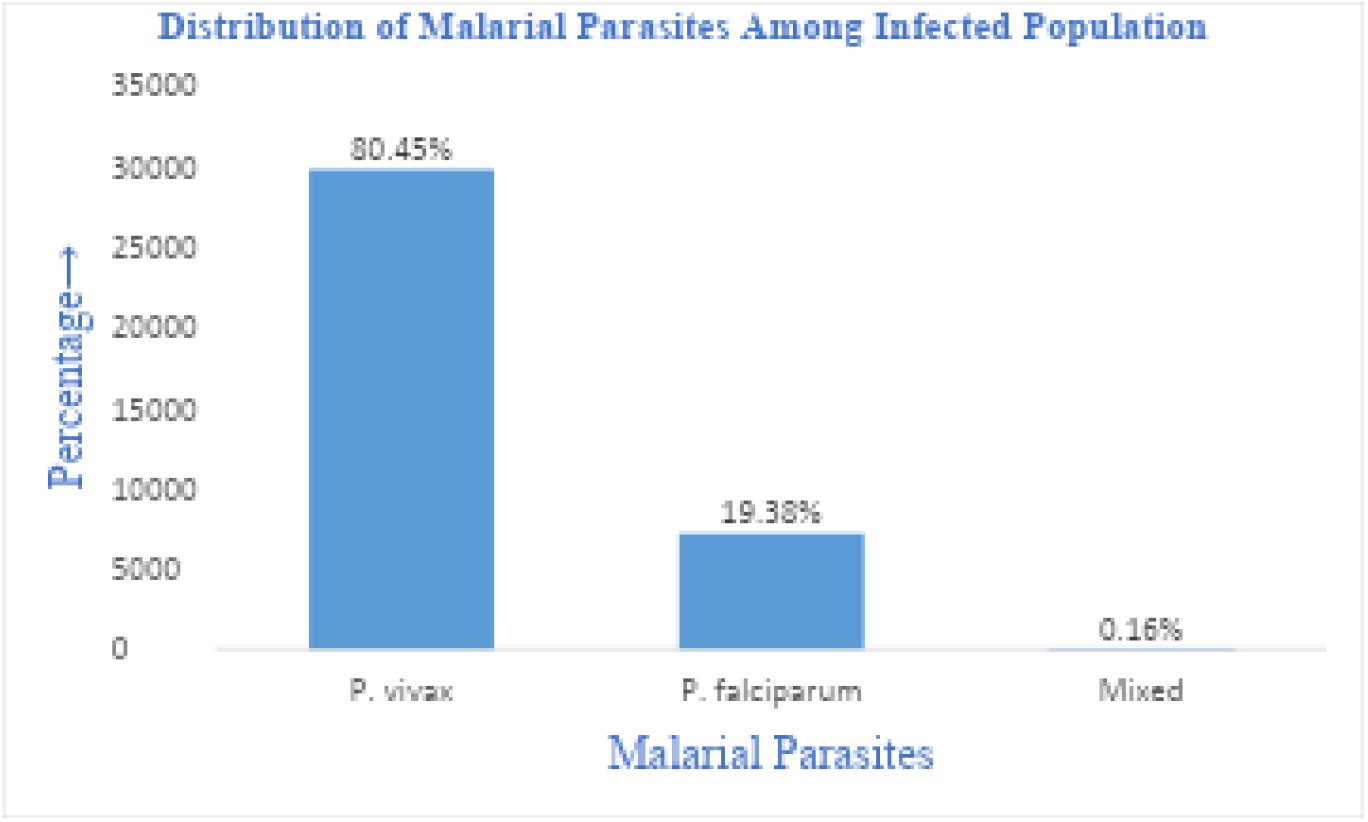
Distribution of Malarial Parasites Among Infected Population

### Overall Sex-wise Distribution of Malarial Parasites

A Chi-square (χ^2^) test of independence was conducted to assess the association between gender (male and female) and the type of malaria parasite (*P. vivax, P. falciparum*, and mixed infections). Among the 36,938 cases, 21,242 were males and 15,696 were females. *P. vivax* accounted for 15,564 male and 14,154 female cases. *P. falciparum* was identified in 5,660 males and 1,500 females, while mixed infections occurred in 18 males and 42 females. At a significance level of 0.05, the critical χ^2^ value is 5.99, and the observed χ^2^ value exceeded this threshold, indicating a statistically significant relationship between gender and malaria species (Table 2) (Figure 2). *P. falciparum* was more prevalent in males, whereas mixed infections were more frequently observed in females.

**Table 2:** Malarial Species Distribution by Gender.

| Species | (Male) | (Female) |
| --- | --- | --- |
| *P. vivax* | 15,564 | 14,154 |
| *P. falciparum* | 5,660 | 1,500 |
| Mixed Infection | 18 | 42 |
| $\chi^2 = 1718.55$; $df = 2$; $\alpha = 5.99$ | | |

**Fig. 2:**
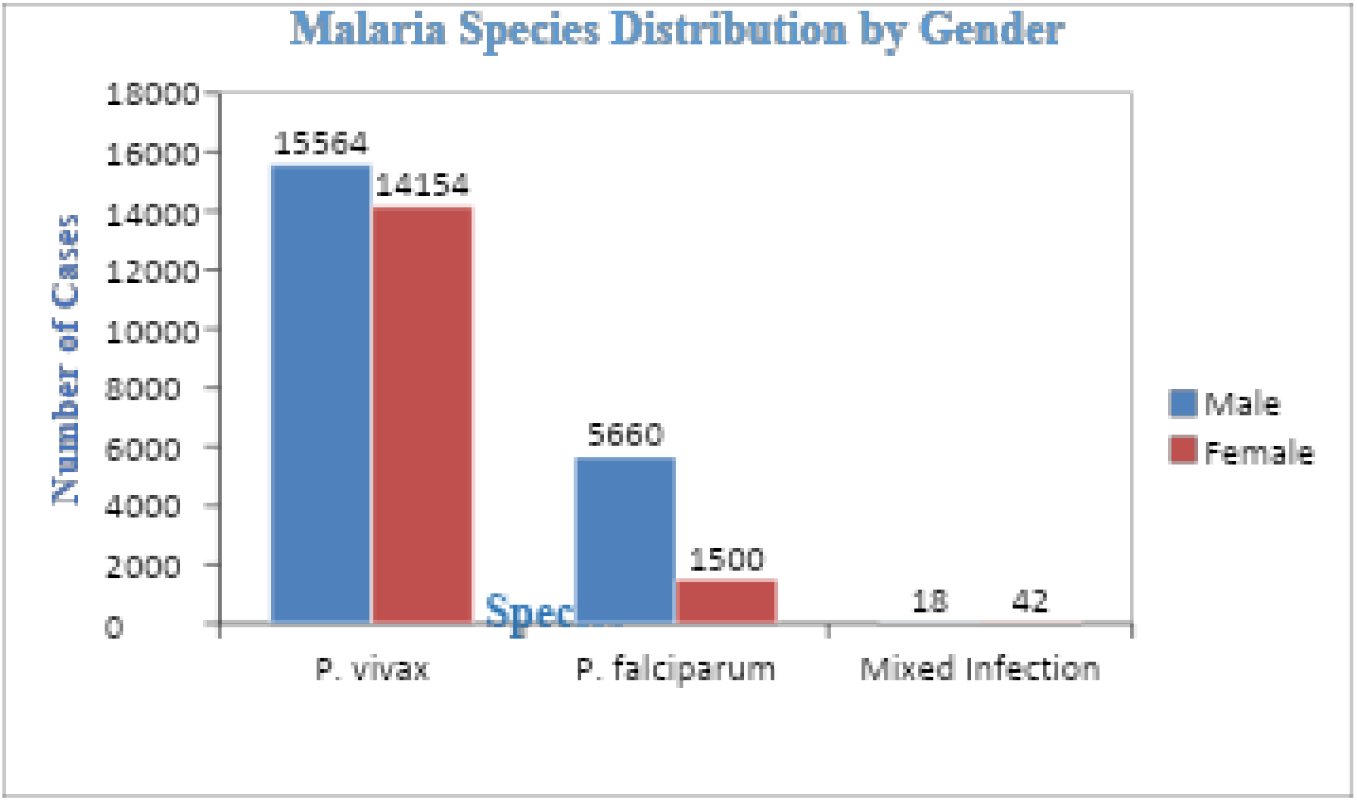
Malarial Species Distribution by Gender

### Age-wise Distribution of Malarial Parasites

Cases were categorized into three age groups: 0–4, 5–14, and 15+ years. In children aged 0–4 years, there were 3,454 *P. vivax*, 188 *P. falciparum*, and 16 mixed infections. In the 5–14 age group, 10,987 cases were *P. vivax*, 106 were *P. falciparum*, and 18 were mixed infections. Among individuals aged 15 years and older, 21,890 cases were *P. vivax*, 240 were *P. falciparum*, and 42 were mixed infections. The p-value was <0.01, indicating a statistically significant correlation between age groups and malaria species. Age significantly influenced the prevalence of different malaria types, with *P. vivax* being the most common species across all age groups (Table 3) (Figure 3).

**Table 3:** Age-wise Prevalence of Malarial Parasites in Among Infected Population.

| Age Group | *P. vivax* | *P. falciparum* | Mixed |
| --- | --- | --- | --- |
| 0-4 | 3454 | 188 | 16 |
| 5-14 | 10987 | 106 | 18 |
| 15+ | 21890 | 240 | 42 |
| $\chi^2 = 401.32$; $df = 4$; $P = \leq 1$ | | | |

**Fig. 3:**
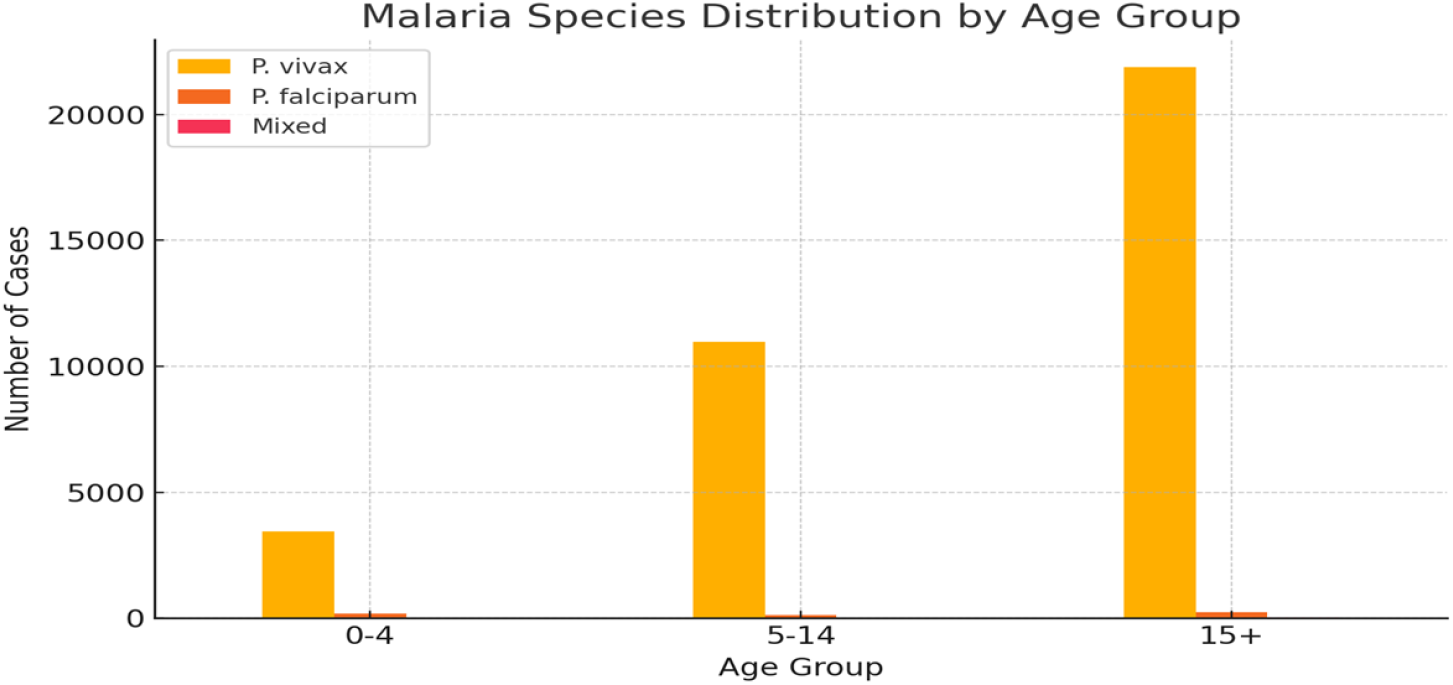
Age-wise Prevalence of Malarial Parasites

### Monthly Distribution of Malarial Parasites

Over the year, 232,779 blood samples were tested for malaria. The highest number of samples were examined in October (28,716; 12.3%), followed by September (26,101; 11.2%) and November (24,392; 10.4%). *P. vivax* peaked in September (5,312; 2.3%), October (6,541; 2.8%), and November (4,464; 1.9%). *P. falciparum* cases were highest in October (436; 1.0%) and November (1,092; 0.4%). Mixed infections were rare, peaking in November (80; 0.03%) and December (61; 0.02%).

Overall, malaria-positive cases exhibited a distinct seasonal pattern, with a marked increase from September to December, following the monsoon season. October recorded the highest positivity rate (7,002; 3.0%), followed by September (5,587; 2.4%) and November (5,636; 2.4%). The Chi-square test confirmed a statistically significant association between malaria species and month, highlighting the strong seasonal influence on malaria prevalence and distribution in the district (Table 4) (Figure 4).

**Table 4:** Month-wise Prevalence of Malarial Parasites.

| Month | Samples tested<br>n (%) | *P. vivax* Positive n<br>(%) | *P.f* Positive n<br>(%) | Both (*P.v* and<br>*P.f*) | Total Positive n<br>(%) |
| --- | --- | --- | --- | --- | --- |
| January | 13,643 (5.8) | 945(0.4) | 203(0.08) | 15 (0.0) | 1,163 (0.5) |
| February | 13164 (5.6) | 691(0.2) | 63 (0.02) | 3 (0.0) | 757(0.3) |
| March | 13,987 (5.9) | 776(0.3) | 41 (0.01) | 24(0.01) | 841(0.3) |
| April | 15,906 (6.8) | 1373(0.5) | 39 (0.02) | 2(0.0) | 1,414(0.6) |
| May | 19764 (8.4) | 2468(1.0) | 196 (0.08) | 3(0.0) | 2,667(1.1) |
| June | 16565 (5.0) | 2213(0.9) | 96 (0.04) | 2(0.0) | 2,311(0.9) |
| July | 17752 (7.6) | 2216(0.9) | 25 (0.01) | 3(0.001) | 2,244(0.9) |
| August | 20326 (8.7) | 2519(1.0) | 44 (0.01) | 2(0.0) | 2,565(1.1) |
| September | 26101 (11.2) | 5312(2.3) | 263 (0.11) | 12(0.005) | 5,587(2.4) |
| October | 28716 (12.3) | 6541(2.8) | 436 (1.0) | 25(0.01) | 7,002(3.0) |
| November | 24392 (10.4) | 4464(1.9) | 1092 (0.4) | 80(0.03) | 5,636(2.4) |
| December | 22463 (9.6) | 3524(1.5) | 839(0.3) | 61(0.02) | 4,424(1.9) |
| $\chi^2 = 2314.40$; df = 22; P = <0.0001 | | | | | |

**Fig. 4:**
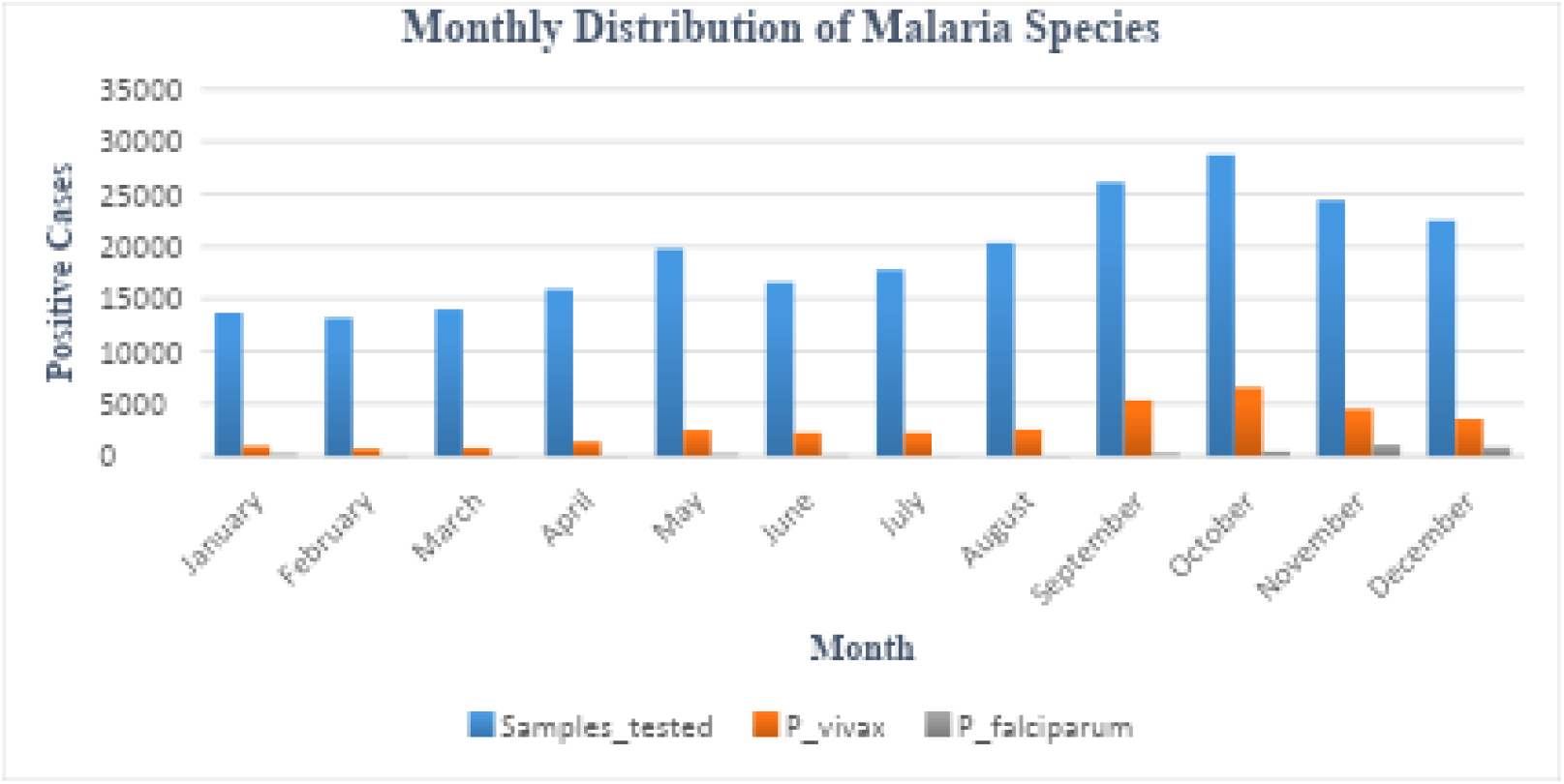
Month-wise Prevalence of Malarial Parasites in Infected Population

## Discussion

The present study assessed the prevalence of malaria in District Dera Ismail Khan from January 2024 to December 2024 and found an overall prevalence of 15.9%, representing a notable increase from the 12.29% reported two years earlier. This rise highlights a growing burden of malaria in the district. *Plasmodium vivax* was identified as the predominant species, followed by *P. falciparum*, while mixed infections were rare (10). These findings are consistent with previous studies in Dera Ismail Khan and other regions of Pakistan, where *P. vivax* has been documented as the major cause of malaria (11). The predominance of *P. vivax* may be attributed to its adaptation to subtropical and temperate regions (12)(13).

Malaria was more prevalent among males (57.5%) compared to females (42.49%), likely due to increased outdoor exposure and occupational activities, as males are more frequently in environments conducive to mosquito bites and are generally less covered than females. Similar gender patterns have been observed in other districts of Pakistan, including Gadap, Buner, Lal Qilla, Charsada, Bannu, Ziarat, Sanjavi, and areas along the Pakistan–Iran border (14).

The seasonal distribution of malaria showed a clear peak in September, October, and November, corresponding with the post-monsoon period, while January and February recorded the lowest incidence. This seasonal trend aligns with previous studies in the district (15) and can be explained by favorable conditions for mosquito breeding, such as increased rainfall and moderate temperatures, which promote vector proliferation (16). Conversely, low temperatures and dry conditions in winter restrict mosquito development and transmission (17). Local irrigation practices may also contribute to the seasonal distribution of malaria cases.

Age-wise analysis indicated that malaria predominantly affected individuals in their middle years before full adulthood, with *P. vivax* remaining the most prevalent species across all age groups. Species-wise prevalence further confirmed this, with 29,718 cases (80.46%) of *P. vivax*, 7,160 cases (19.39%) of *P. falciparum*, and only 60 cases (0.16%) of mixed infections. Males were more affected by *P. falciparum*, while mixed infections were slightly more common in females. Monthly analyses reinforced the distinct seasonal pattern, with the highest malaria-positive cases occurring from September to December, following the monsoon season.

The findings of this study underscore that *P. vivax* is the dominant malaria species in Dera Ismail Khan, malaria incidence is higher in males than females, and middle-aged individuals are most affected. The study also confirms a clear seasonal pattern of malaria transmission, with peak prevalence in the post-monsoon months. These insights are critical for guiding targeted malaria control and prevention strategies in the district.

Despite the strengths of this study, several limitations must be acknowledged. First, data were obtained from secondary sources (District Health Office), which may limit verification and affect the accuracy, consistency, and completeness of records. Second, only microscopically-confirmed cases were analyzed, potentially overlooking subclinical or asymptomatic infections. Third, while age, gender, and basic clinical characteristics were included, other important factors such as socio-economic status were not considered. Additionally, the cross-sectional design restricts causal inferences and limits long-term assessment of seasonal trends. Finally, molecular identification of Plasmodium species and drug resistance profiling were beyond the scope of this study, which may limit the accuracy of species-specific diagnostics and treatment insights.

Overall, the study provides important epidemiological evidence regarding the prevalence, demographic patterns, and seasonal distribution of malaria in Dera Ismail Khan, which can inform more effective, locally tailored public health interventions.

## Data Availability

Data can be accessed from the District Health Office (DHO) Dera Ismail Khan

## Acknowledgment

The authors wish to extend their sincere gratitude to the District Health Office (DHO) of Dera Ismail Khan for their invaluable assistance and for providing the essential information that made this research possible. We would also like to express our deep appreciation to Abdus Sami for his continuous encouragement, insightful suggestions, and guidance throughout the process of writing this article. His support was instrumental in shaping both the direction and quality of this work.

